# A novel approach for estimating vaccine efficacy for infections with multiple outcomes: application to a COVID-19 vaccine trial

**DOI:** 10.1101/2023.03.02.23286698

**Authors:** Lucy R Williams, Merryn Voysey, Andrew J Pollard, Nicholas C Grassly

## Abstract

Vaccines can provide protection against infection or limit disease progression and severity. Vaccine efficacy (VE) is typically evaluated independently for different outcomes, but this can cause biased estimates of VE. We propose a new analytical framework based on a model of disease progression for VE estimation for infections with multiple possible outcomes of infection: Joint analysis of multiple outcomes in vaccine efficacy trials (JAMOVET). JAMOVET is a Bayesian hierarchical regression model that controls for biases and can evaluate covariates for VE, the risk of infection, and the probability of progression. We applied JAMOVET to simulated data, and data from COV002 (NCT04400838), a phase 2/3 trial of ChAdOx1 nCoV-19 (AZD1222) vaccine. Simulations showed that biases are corrected by explicitly modelling disease progression and imperfect test characteristics. JAMOVET estimated ChAdOx1 nCoV-19 VE against infection (*VE*_*in*_) at 49% (95% CI 37-59) and progression to symptoms (*VE*_*pr*_) at 44% (95% CI 27-58). This implies a VE against symptomatic infection of 72% (95% CI 63-80), consistent with published trial estimates. *VE*_*in*_ decreased with age while *VE*_*pr*_ increased with age. JAMOVET is a powerful tool for evaluating diseases with multiple dependent outcomes and can be used to adjust for biases and identify predictors of key outcomes.

## 1 Introduction

The evaluation of vaccine efficacy and effectiveness in clinical trials and post-licensure studies needs to be carefully designed and interpreted to be statistically efficient and unbiased – thereby providing maximum value of information for policy decisions and scientific understanding. Infectious diseases have several features that undermine these goals, including heterogeneity in risk of infection and disease progression, rapid dynamics, and outcomes that depend on the vaccination status of contacts. Mathematical and statistical modelling can determine how these features affect the validity of vaccine efficacy (VE) estimates^1-4^ and is increasingly important for study design, power calculations and analysis of results^5^. Insights from models support innovative trial designs that can make the difference between success or failure in the evaluation of VE^6^.

Infection typically results in multiple dependent outcomes, reflecting disease natural history (e.g., different degrees of disease severity) or different disease presentations (e.g., symptom groupings) (Figure 1). Estimates of VE vary when measured against these different outcomes. For example, VE of licensed rotavirus vaccines is usually lower when measured in studies of less versus more severe disease^7^. In some cases, estimated VE against intermediate outcomes such as asymptomatic infection may be negative – not because vaccination increases infection but because it prevents progression to symptomatic or more severe disease^8^.

**Figure 1.**
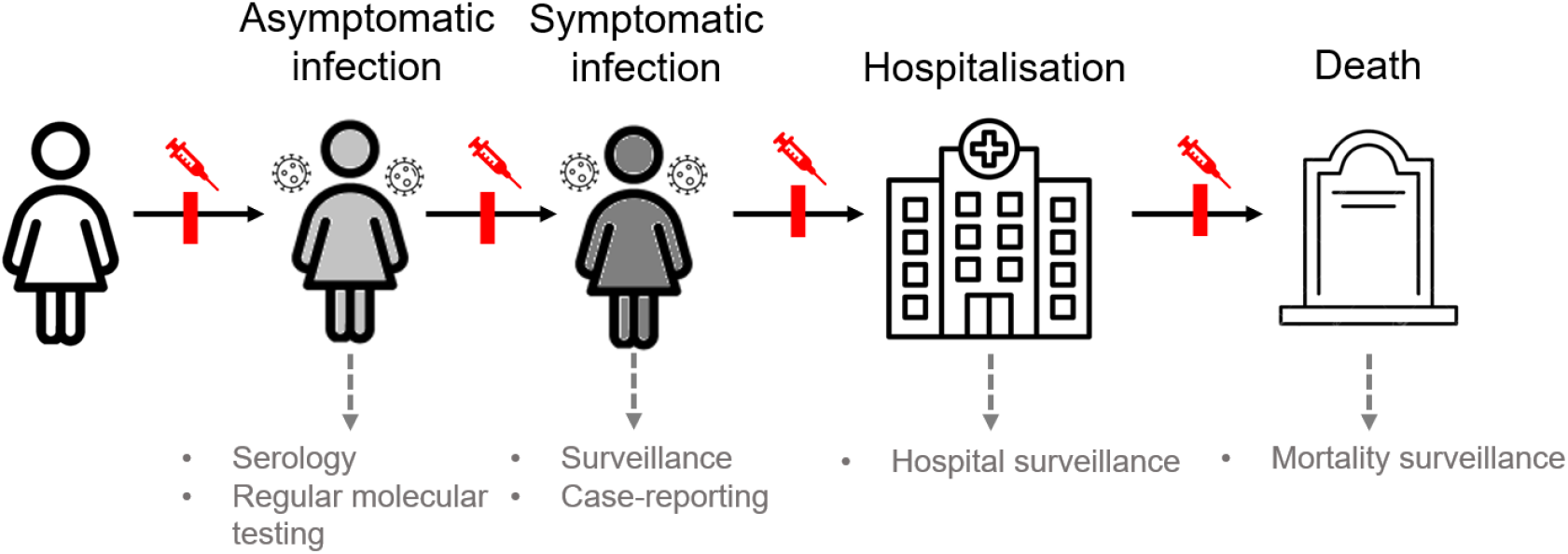
Vaccine effects and detection methods at different stages of infection and disease progression.

Typically, in clinical trial analyses, VE is evaluated from the relative risk of each outcome separately, and common statistical approaches include Poisson and Cox proportional hazards regression^9^. In addition to the challenges in interpretation of VE against intermediate outcomes, independent analysis of each outcome is susceptible to biases. For example, a differential detection bias arises if less severe outcomes are less likely to be detected and vaccination prevents disease progression, resulting in an overestimation of the overall VE against infection^10^. This is because the act of the vaccine in preventing an infection from progressing from one stage to another results in that infection becoming less likely to be detected, contributing to apparent efficacy against infection.

Other important biases are also rarely incorporated into classical clinical trial analyses. For example, the accumulation of false positives in both trial arms as a result of using a test with imperfect specificity can lead to underestimation of VE^11,12^. This bias can be particularly impactful in a setting where the number of false positives is large in comparison to the total number of cases included in the analysis, which can occur if testing is performed frequently, the force of infection is low, and if test sensitivity is also low. This impact of false positives is not usually adjusted for in clinical trial analyses as the direction of potential bias is towards an under- rather than over-estimation of VE, thus the resultant analysis is conservative, which is usually preferred.

Multistate Markov models are a powerful and flexible tool, typically used to analyse longitudinal data on disease progression in population cohorts^13,14^. However, they have not been used in the context of vaccine efficacy evaluation in clinical trials^14-16^. This is surprising given the progressive nature of many vaccine-preventable diseases, as the multistate model approach lends itself to identifying the stage in the disease process at which the vaccine is effective.

In this paper, we propose a new analytical framework for the integrated analysis of VE for infections with multiple disease outcomes that is an adaptation of the multistate model framework: Joint analysis of multiple outcomes in vaccine efficacy trials (JAMOVET). JAMOVET can control for biases and evaluate predictors of VE, the risk of infection, and the probability of progressing to each disease stage. We first validate the model using simulated data and then apply it to data from a phase 3 COVID-19 vaccine trial.

## 2 Methods

### 2.1 Model Framework

JAMOVET can be described as a hierarchical multivariable generalized regression model. It models multiple progressive stages of a disease and may have covariates for the parameters describing disease progression and VE. The basic model framework can be illustrated by first considering the simple case of a clinical trial of a vaccine against an infection that may be symptomatic or asymptomatic. We adopt a regression framework such that the number of symptomatic infections experienced by individual *i* in a given time period (e.g., length of follow-up) is assumed to follow a Poisson distribution

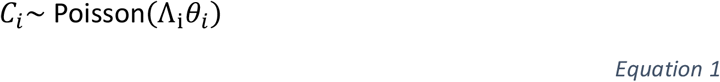

where Λ_i_ and *θ*_*i*_ are the cumulative hazard or force of infection (FOI) and the probability of symptoms for that individual, respectively.

With perfect test sensitivity and specificity, the number of asymptomatic infections is given by

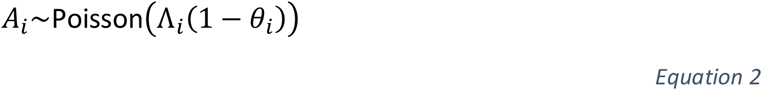

The cumulative FOI is estimated in a log-linear regression model with an offset for follow-up time and vaccination status as an indicator variable (0=placebo, 1=vaccine).

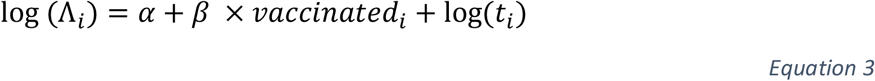

where *α* is the log of the average FOI in the placebo arm during the follow-up period and *β* is the log relative risk of infection as a result of vaccination. The VE against infection, *VE*_*in*_, can be calculated as

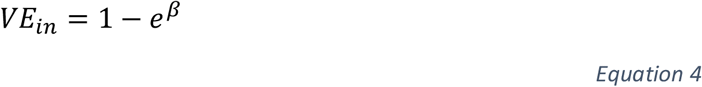

The probability of symptoms is estimated using logistic regression and depends on vaccination status.

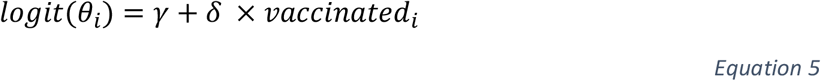

Where *γ* is the log odds of symptom development in the placebo arm and *δ* is the log odds ratio of symptom development in the vaccine compared to the placebo arm. We estimate VE against progression, *VE*_*pr*_ from the relative risk (RR) of progression in the vaccine compared with the control arm (*VE*_*pr*_ = 1 − *RR*, rather than using the odds ratio 1 − *e*^*δ*^)^17^.

### 2.2 Model extensions

JAMOVET can be extended to account for additional stages of severity (Supplementary Methods), imperfect test characteristics and differential detection of different outcomes, such as a lower probability of detecting asymptomatic infections compared to symptomatic. For example, consider the case where regular samples are taken from asymptomatic individuals to test for the presence of infection (as was done in some COVID-19 vaccine trials). If *σ* is the probability of detecting an asymptomatic infection, *m*_*i*_ is the number of tests taken by individual *i*, and *q* is the test specificity for asymptomatic infections (ignoring false positive symptomatic infections that are likely to be rare), then the number of asymptomatic infections is given by:

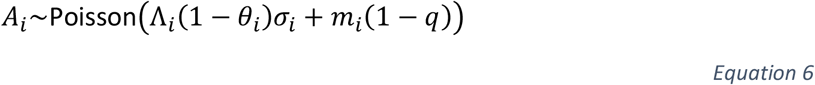

The model can also be extended to model covariates on the force of infection and probability of symptoms by including these terms in their respective regression equations. Interaction terms on the vaccination indicator variables also allows for covariates on *VE*_*in*_ and *VE*_*pr*_ to be modelled (Supplementary Methods).

### 2.3 Application to simulated data

To demonstrate the accuracy of the approach we applied JAMOVET to four simulated datasets. All datasets had a sample size of 10,000, similar to the COV002 ChAdOx1 nCoV-19 (AZD1222) vaccine trial, and participants were followed up for one year and censored after infection. For all participants, the true Λ = 10% per year, *θ* = 0.6, *VEi*_*n*_ = 50% and *VE*_*pr*_= 50%. This corresponds to a true VE against symptomatic infection (*VE*_*sym*_ = 1 − (1 − *VE*_*in*_)(1 − *VE*_*pr*_)) of 75% and VE against asymptomatic infection (*VE*_*asym*_ = 1 − ((1 − *θ*)(1 − *VE*_*in*_) + *θ*(1 − *VE*_*in*_)*VE*_*pr*_)/(1 − *θ*)) of 12.5%^18^.

In dataset 1, we introduced no bias from imperfect test characteristics or differential detection (Table 1). In dataset 2, we simulated false positives using a test with specificity of 99.9%, with weekly asymptomatic testing. In dataset 3, we made asymptomatic infections 50% less likely to be detected than symptomatic infections (representative of either imperfect test sensitivity or incomplete adherence to testing). Finally, in dataset 4, we assumed weekly testing with a 99.9% specific, 100% sensitive test, with 50% adherence to asymptomatic testing. The asymptomatic infection positive predictive values across trial arms and datasets varied between 38% (95% CI 34-43) and 100% (95% CI 100-100) (Supplementary Table 1). We applied JAMOVET to each dataset, adjusting for biases where appropriate, to estimate the FOI, *p*_*s*_, *VE*_*in*_, *VE*_*pr*_, *VE*_*sym*_ and *VE*_*asym*_. We compared the results to a separate analytical approach where *VE*_*in*_, *VE*_*sym*_ and *VE*_*asym*_ were estimated individually using standard Poisson regression with an offset for time. Both JAMOVET and the standard separate analysis were conducted without covariates on any parameters.

**Table 1.** Parameter values in simulated datasets.

| Scenario | Test specificity ($q$, %) | Prob. asymptomatic infection detection ($\sigma$, %) |
| --- | --- | --- |
| 1 | 100.0 | 100.0 |
| 2 | 99.9 | 100.0 |
| 3 | 100.0 | 50.0 |
| 4* | 99.9 | 50.0 |

\*Modelled as incomplete adherence to testing rather than imperfect test sensitivity (50% detection of true positives and false positives)

**Table 2.** Vaccine efficacy estimates using separate analysis and JAMOVET analysis on simulated datasets.

| True parameter value (%)<sup>1</sup> |  | Model estimated value (%) |  |  |  |  |  |  |  |
| --- | --- | --- | --- | --- | --- | --- | --- | --- | --- |
|  |  | Scenario 1<br>Specificity and prob.<br>asymptomatic infection<br>detection = 100% |  | Scenario 2<br>Specificity = 99.9% |  | Scenario 3<br>Prob. asymptomatic infection<br>detection = 50% |  | Scenario 4<br>Specificity = 99.9% and prob.<br>asymptomatic infection<br>detection = 50%<sup>3</sup> |  |
|  |  | Separate | JAMOVET<sup>2</sup> | Separate | JAMOVET<sup>2</sup> | Separate | JAMOVET<sup>2</sup> | Separate | JAMOVET<sup>2</sup> |
| $VE_{in}$ | 50 | 49.51<br>(41.28-56.85) | 49.90<br>(41.84-57.43) | 32.40<br>(24.35-40.17) | 49.73<br>(38.23-60.44) | 58.83<br>(51.07-66.07) | 49.86<br>(40.18-59.45) | 44.19<br>(34.76-51.91) | 49.15<br>(34.29-62.55) |
| $VE_{pr}$ | 50 | -<br>(-) | 49.89<br>(39.02-59.37) | -<br>(-) | 49.21<br>(34.64-61.23) | -<br>(-) | 49.72<br>(35.87-60.52) | -<br>(-) | 49.77<br>(32.63-63.07) |
| $VE_{sym}$ | 75 | 74.41<br>(67.89-80.46) | 74.90<br>(68.35-80.84) | 74.30<br>(67.22-81.12) | 74.75<br>(67.75-81.57) | 74.42<br>(67.81-80.43) | 74.91<br>(68.27-80.85) | 74.33<br>(67.49-81.04) | 74.84<br>(68.17-81.52) |
| $VE_{asym}$ | 12.5 | 10.76<br>(-9.15-27.59) | 12.14<br>(-7.01-28.53) | 4.43<br>(-8.42-16.69) | 10.09<br>(-21.39-37.44) | 8.96<br>(-18.91-32.40) | 11.72<br>(-14.88-34.99) | 3.53<br>(-17.32-20.71) | 7.67<br>(-38.19-41.34) |

<sup>1</sup> $VE_{in}$ and $VE_{pr}$ were parameters in the models. $VE_{sym}$ and $VE_{asym}$ were calculated from these parameters: $VE_{sym} = (1 - VE_{in})(1 - VE_{pr})$ and $VE_{asym} = \frac{(1-\theta)(1-VE_{in})+\theta(1-VE_{in})VE_{pr}}{(1-\theta)}$ , where $\theta$ is the probability of symptoms in the unvaccinated.
<sup>2</sup> Joint analysis of multiple outcomes in vaccine efficacy trials with bias-correction applied.
<sup>3</sup> Modelled as incomplete adherence to testing rather than imperfect test sensitivity (50% detection of true positives and false positives)

### 2.4 Application to COV002 phase III trial

We then applied JAMOVET to data from the University of Oxford sponsored phase 2/3 clinical trial, COV002 (NCT04400838). COV002 is a single-blind, multicentre, randomised phase 2/3 trial assessing the safety and efficacy of the SARS-CoV-2 ChAdOx1 nCoV-19 (AZD1222) vaccine. The full protocol and statistical analysis plan have been previously published in detail^19-22^. Participants were adults aged 18 years and older, enrolled at 19 study sites across the United Kingdom to either the phase 2 (immunogenicity) or phase 3 (efficacy) cohort. Enrolment began on May 28, 2020 and targeted participants in occupations with potentially high SARS-CoV-2 exposure, such as healthcare workers (HCWs). Participants were randomly assigned to receive the ChAdOx1 nCoV-19 vaccine or a control vaccine. In this analysis we have data until the February 25^th^, 2021 data cut, thus most infections were due to alpha or earlier variants.

Symptomatic infections were detected through responsive symptomatic reporting and RT-PCR testing, while asymptomatic and pre-symptomatic infections were detected through regular asymptomatic RT-PCR swab testing. Any infections detected through asymptomatic swab collection that became symptomatic (i.e., pre-symptomatic infections) were classified as a single event of a symptomatic case.

We used the baseline-seronegative subset of the phase III (efficacy) cohort for this analysis and excluded participants who did not receive two doses of vaccine or the control, or who experienced a SARS-CoV-2 infection before two weeks following their second dose (n=9139). As study participants were allowed to unblind once SARS-CoV-2 vaccines became publicly available, we censored after i) SARS-CoV-2 infection, ii) unblinding, and iii) February 25^th^, 2021, whichever was earliest.

Test specificity was based on published estimates for a low prevalence setting (99.945%^23^) and we made the simplifying assumption that all symptomatic infections were detected. The probability of asymptomatic infection detection was estimated for each individual as a function of their adherence to weekly asymptomatic testing and PCR test sensitivity to asymptomatic SARS-CoV-2 infections over time since infection^24^ (Supplementary Methods).

Yearly FOI, probability of symptoms, *VE*_*in*_ and *VE*_*pr*_ were directly estimated in the model, with *VE*_*sym*_ and *VE*_*asym*_ calculated from *VE*_*in*_ and *VE*_*pr*_, as above^10,25^.

We adjusted for covariates on the FOI, probability of symptoms (PS), *VE*_*in*_ and *VE*_*pr*_. Candidate covariates were:

- Age
- Sex
- Body mass index (BMI)
- Ethnicity
- HCW status
- Comorbidities

Both categorical and continuous covariates were considered (Supplementary Table 2).

Variable selection was based on model fit, measured by leave one out (loo) cross validation using the loo R package^26^. A univariable model was fit for each variable for each parameter (FOI/PS/*VE*_*in*_/*VE*_*pr*_) and those that improved the model fit compared to a model without any covariates were selected for a full model. Any variables that improved the fit for *VE*_*in*_were also selected for FOI and any that improved the fit for *VE*_*pr*_ were also selected for the probability of symptoms, PS. Variables were then removed from the full model by backwards selection, incrementally removing the variable with the smallest effect until the model fit no longer improved. The type of variable (categorical/continuous) was selected as the one that gave the best model fit.

We compared the results of this final model with the results of an unadjusted model, and models that adjust only for i) imperfect test specificity, ii) differential detection, and iii) covariates.

### 2.5 Model fitting and parameter estimation

We implemented all models using Hamiltonian Monte Carlo algorithms, as implemented in the Rstan package^27^ in the R statistical environment^28^. All intercept parameter priors were given a Cauchy distribution with centre 0 and scale 10^29^. Other regression coefficients were given Cauchy priors with centre 0 and scale 2.5. Continuous predictors were scaled to have mean 0 and standard deviation 0.5, while categorical predictors were converted to dummy binary predictors and were shifted to have a mean of 0 and differ by 1 in their lower and upper conditions. Sensitivity analysis using uniform, non-informative priors on all regression coefficients, bounded between -50 and 50 showed consistent results, so these priors were used for the simulation models. All analyses were run with 4 chains, with a warmup period of 1000 iterations and a total of 2000 iterations. Convergence was diagnosed based on the 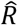 convergence diagnostic (all 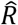 values were below 1.05 for all parameters) and trace plots that indicated good mixing. Code is available at: https://github.com/lucyrose96/JAMOVET.

## 3 Results

### 3.1 Simulation results

Applied to simulated data with perfect test characteristics, the separate models and JAMOVET produced accurate and comparable results (Table 1, Scenario 1). For data simulated with imperfect test specificity for asymptomatic infections (Table 1, Scenarios 2 & 4), the separate analysis underestimated *VE*_*in*_ (true = 50%, Scenario 2 estimated = 32.40% [95% CI 24.35 to 56.88], Scenario 4 estimated = 44.19 [95% CI 34.76-51.91]), while JAMOVET with bias adjustment accurately estimated all VE outcomes (Figure 2). When asymptomatic infections were 50% less likely to be detected than symptomatic infections (Table 1, Scenario 3), the separate analytical approach overestimated *VE*_*in*_ by an absolute difference of 8.8% (95% CI 1.1 to 16.1), while JAMOVET gave an unbiased estimate (absolute difference 0.1% [95% CI -9.8 to 9.5]).

**Figure 2.**
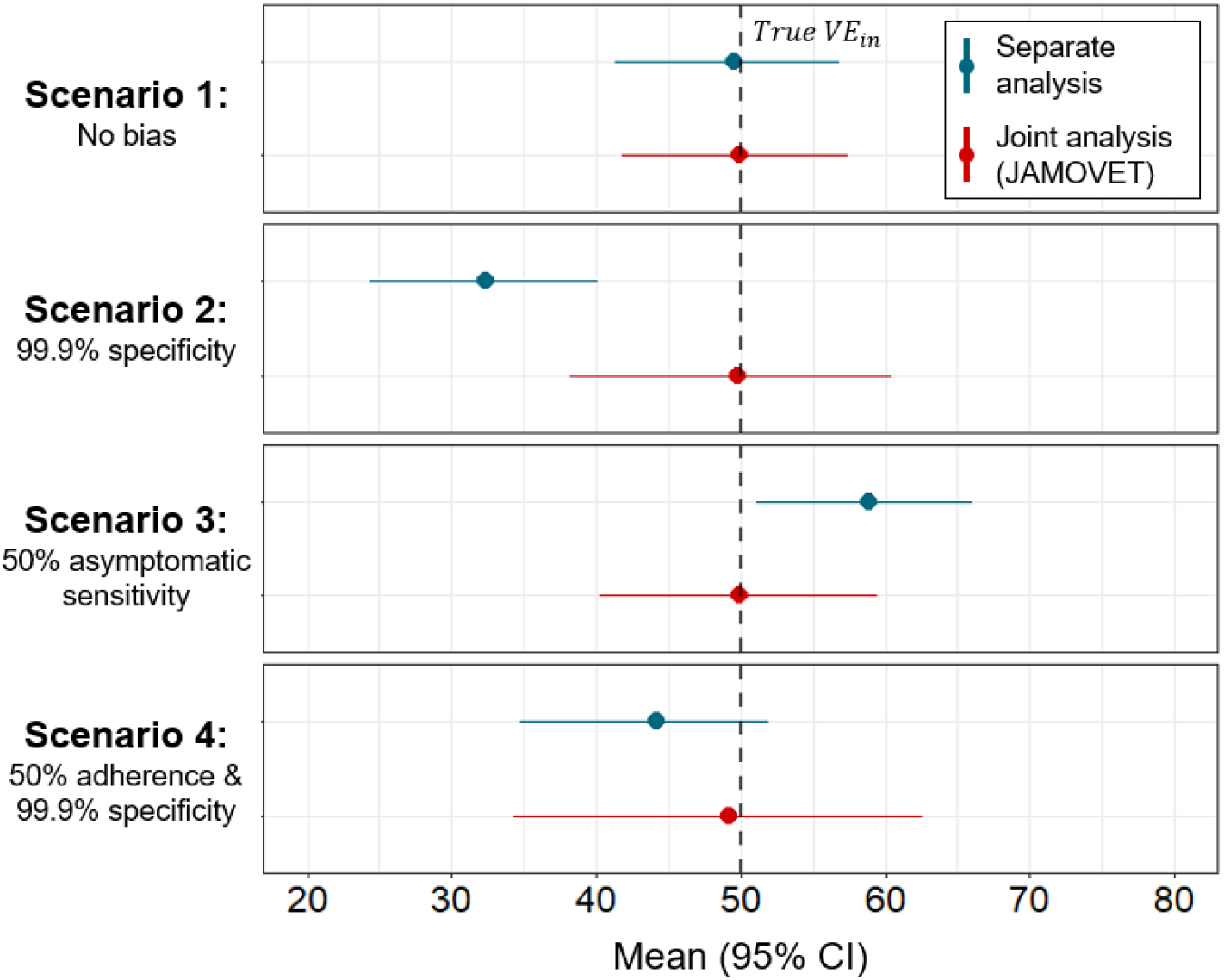
Vaccine efficacy against infection (VE_in_) estimates for separate analysis and JAMOVET analysis with bias adjustment.

### 3.2 COV002 results

298 (3.43%) and 291 (3.35%) participants experienced symptomatic and asymptomatic SARS-CoV-2 infections before 25th February 2021, respectively. Most symptomatic cases were of mild or moderate severity, with only three participants requiring hospitalisation as a result of SARS-CoV-2 infection and two classifying as severe (Supplementary Table 3). The median proportion of expected tests returned was 88.0%. Combined with known test sensitivity, this gave a median estimated probability of asymptomatic infection detection of 78.8% (IQR 70.3 to 83.4).

Applying JAMOVET to COV002 without bias adjustment gave almost identical VE estimates to those reported from the trial (Table 3)^30^. It also estimated the yearly FOI at 31% (95% CI 28-34) and the probability of symptoms at 59% (95% CI 54-64). Adjusting for false negatives and incomplete adherence to regular asymptomatic testing decreased the estimated *VE*_*in*_ by an absolute difference of 4% and increased the estimated *VE*_*pr*_ by 6%. Accounting for imperfect test specificity made a greater difference to the model estimates, increasing estimated *VE*_*in*_ by 5% and decreasing estimated *VE*_*pr*_ by 6%. As expected, adjusting for a lower probability of detecting asymptomatic infections decreased the estimated probability of symptoms, while accounting for false positive asymptomatic infections increased this parameter.

**Table 3.** JAMOVET estimates for COV002 analysis population with bias and covariate adjustments.

|  | Trial-reported results <sup>30</sup> | Joint model estimated mean (95% CI) |  |  |  |  |
| --- | --- | --- | --- | --- | --- | --- |
|  |  | No adjustment | Sensitivity<sup>1</sup> only | Specificity only | Covariates only | All adjustments |
| $VE_{in}$ (%) | 51 (41-59) | 51 (42, 59) | 47 (37, 56) | 56 (47, 64) | 49 (40, 58) | **49 (37-59)** |
| $VE_{pr}$ (%) | - | 41 (29, 53) | 47 (33, 59) | 35 (20, 48) | 45 (31, 57) | **44 (27-58)** |
| $VE_{sym}$ (%) | 72 (63-79) | 71 (63, 78) | 72 (63, 79) | 71 (63, 79) | 72 (63, 80) | **72 (63-80)** |
| $VE_{asym}$ (%) | 15 (-12-35) | 22 (2, 39) | 22 (2, 39) | 28 (4, 48) | 11 (-18, 32) | **13 (-88-39)** |
| Yearly FOI (%) | - | 31 (28, 34) | 37 (33, 40) | 28 (25, 31) | 30 (27, 33) | **31 (27-35)** |
| Prob. Symptoms (%) | - | 59 (54, 64) | 50 (45, 55) | 64 (59, 69) | 56 (51, 61) | **54 (48-60)** |

$VE_{in}$ = Vaccine efficacy against infection; $VE_{pr}$ = vaccine efficacy against progression to symptoms, $VE_{sym}$ = vaccine efficacy against symptomatic infection, $VE_{asym}$ = vaccine efficacy against asymptomatic infection.
<sup>1</sup>Accounts for imperfect PCR test sensitivity and incomplete adherence to regular asymptomatic testing.

Several variables were identified as predictive of one of the four key parameters. This is shown in Figure 3, where the reference is an example subgroup with the following characteristics: 30 years, non-healthcare worker, not obese, and white ethnicity. Other example subgroups are shown in black to show how modifying one of the characteristics changes each estimate. The FOI decreased with age, while HCWs, especially those seeing COVID-19 patients had a greater risk of SARS-CoV-2 infection. The probability of developing symptoms was slightly higher in HCWs who saw COVID-19 patients and people classifying as obese (BMI ≥30) or identifying as non-white ethnicity. While *VE*_*in*_ decreased with age (reference age 30 years = 58% [95% CI 45 to 69] vs reference age 60 years = 39% [95% CI 15 to 58]), *VE*_*pr*_ increased with age (reference age 30 years = 42% [95% CI 25 to 58] vs reference age = 60 years 62% [95% CI 38 to 78]), such that the *VE*_*sym*_ was approximately the same across age groups (reference age 30 years 76% [95% CI 66 to 84] vs reference age 60 years 84% [95% CI 73 to 84]).

**Figure 3.**
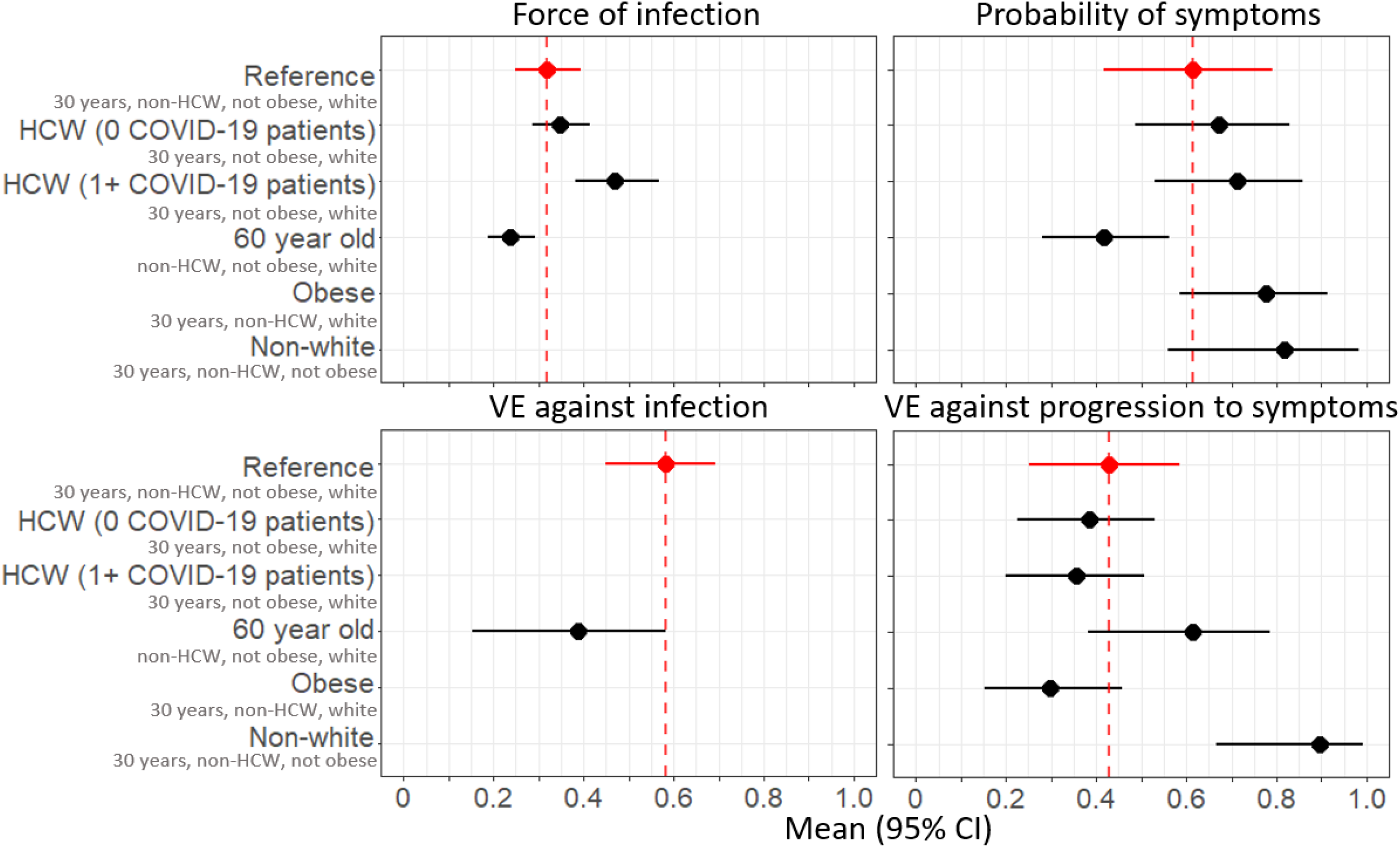
Absolute estimates with 95% credible intervals for each model parameter for example subgroups in the COV002 trial, using the final model (adjusting for sensitivity, specificity, adherence to testing and covariates). Reference = aged 30 years, non-healthcare worker, not obese, white. All other estimates are example subgroups with the reference characteristics and a single characteristic modified.

Overall, adjustment for these covariates and the biases lead to slightly lower *VE*_*in*_ estimates than reported during the trial, (49% [95% CI 37-59] vs 51% [95% CI 41-59] for JAMOVET vs published estimates respectively) but gave consistent results for *VE*_*sym*_ and *VE*_*asym*_. The JAMOVET model also estimated FOI at 31% (95% CI 27 to 35) per year and the probability of symptoms at 54% (95% CI 48 to 60) for the trial population as a whole.

## 4 Discussion

In this paper we have demonstrated the utility and benefits of JAMOVET for the analysis of two dependent VE outcomes (symptomatic and asymptomatic infection). We have shown its accuracy in estimating VE in the presence of strong biases and demonstrated the additional benefits of evaluating socio-demographic predictors of the risk of infection, progression, and VE against each of these outcomes. It therefore provides an integrated, unbiased analytical approach that has considerable advantages over classical separate analyses.

Our simulations showed the large biases that can be corrected using JAMOVET. In our simulation scenarios, the separate analysis approach underestimated *VE*_*in*_ by an absolute difference of 17.6% (95% CI -9.8 to 25.65) when test specificity was imperfect; when there was differential detection of infections based on symptom status, *VE*_*in*_ was overestimated by 8.8% (95% CI 1.1 to 16.1). JAMOVET accurately estimates all VE outcomes in the presence of such biases because it explicitly models the accumulation of false positives and differences in probabilities of detection of different outcomes.

Applied to the COV002 data, the effects of the bias adjustments were relatively small. For example, adjusting for specificity increased the estimated *VE*_*in*_ by 6% while adjusting for differential detection decreased it by 4%. The modest effect of the specificity adjustment is likely because the assumed test specificity was high (99.945%^31^) - we did not account for any potential cross contamination, which could cause additional false positives^32^. When including both covariates and bias adjustment, each counteracted the other (differential detection giving an upwards bias for *VE*_*in*_ and imperfect test specificity giving a downwards bias). Overall, adjusting for biases and covariates, JAMOVET gave a slightly lower estimate for *VE*_*in*_ than reported in the trial^30^, but all other VE estimates were consistent (importantly, JAMOVET *VE*_*sym*_ = 72% [95% CI 63-80], trial-reported *VE*_*sym*_ = 72% [95% CI 63-79], demonstrating robustness to bias-adjustment in this case.

Several predictors of the risk of SARS-CoV-2 infection, probability of progressing to symptoms, and VE were identified for the COV002 data. We identified age as the only predictor of *VE*_*in*_, with participants aged 60 years estimated as having a *VE*_*in*_ a third lower than participants aged 30 years. This is supported by vaccine effectiveness research on the Pfizer-BioNTech (BNT162b2) and the Oxford-AstraZeneca COVID-19 vaccines^33^, in which a greater reduction in risk of post-vaccination infection was found in participants aged 55 years or younger than in those older than 55 years.

However, we also showed that while vaccinated older participants had poorer protection against infection, their risk of symptomatic COVID-19 was approximately the same as younger participants because they showed greater vaccine-induced protection against progression to symptoms. This is supported by vaccine effectiveness research by Antonelli *et al*.^34^, who found a greater reduction in risk of symptom development in the over 60s than in any other age group. It may reflect the remodelling of the immune system with age, with impaired antibody protection but maintenance of an effective T cell response^35,36^. We also identified non-white ethnicity as a predictor of higher *VE*_*pr*_. Due to a small proportion of participants enrolled from ethnic minority groups, we did not have the statistical power to examine this result to a greater resolution, so this may be a target area for further research.

As has been previously shown, HCWs who saw COVID-19 patients and people of younger age had a higher risk of SARS-CoV-2 infection ^34,37,38^ compared with non-healthcare workers and people of older age. Our results and previous work suggest that obesity and non-white ethnicity are associated with more severe COVID-19 outcomes^39,40^. We also found a higher probability of symptoms in HCWs, particularly those that saw COVID-19 patients. High viral dose, likely experienced by many HCWs, has been postulated to be associated with increased symptom severity^41,42^. However, evidence is limited and moreover, personal protective equipment became widespread later in the pandemic. An alternative explanation is that, despite our adjustment for adherence to weekly asymptomatic testing, there may still be some residual differential detection bias, leading subgroups with poorer weekly asymptomatic testing adherence (such as HCWs and younger age groups]) to have inaccurately high estimates for the probability of symptoms.

Bias adjustment for imperfect test sensitivity and specificity required prior estimates of the probability of detecting each type of outcome and the rate of accumulation of false positives (we could not estimate these parameters within the model). Test specificity is usually known, but the number of false positives arising from contamination within the lab is more difficult to quantify, and the relative probabilities of detecting each outcome may also be difficult to estimate. In our analysis we had good literature on PCR sensitivity for asymptomatic infections and on the adherence to regular testing, which allowed us to estimate an individual-based probability of asymptomatic infection detection. Even so there is a risk of residual bias resulting from unadjusted differential detection.

We have only applied our estimation framework to a model of disease progression with two stages. However, the model can be extended to multiple stages including more severe outcomes or alternative disease pathways (Supplementary Methods). In addition, the application of our model to COV002 was limited by the sample size, as we were not able to evaluate all predictors at their smallest category resolution or additional levels of severity beyond symptoms. If applied to a larger clinical trial or vaccine effectiveness study, VE against progression to more severe outcomes could be estimated. For example, this framework could be applied to cohort studies of vaccine effectiveness to estimate VE against progression to hospitalisation, admission to intensive care and death. In addition, JAMOVET can also be used to investigate immune correlates of protection (CoPs), through their mediating effects on VE against infection and disease progression.

There are many benefits of using a Bayesian approach, implemented in Rstan, for our analytical framework. Firstly, it provides posterior probability distributions and 95% credible intervals for model parameters, including VE, which have a more natural interpretation than frequentist maximum likelihood confidence intervals. Secondly, the Bayesian framework provides a natural inferential framework for complex hierarchical models that allows the incorporation of prior knowledge in the form of prior probability distributions for parameters. This prior knowledge may come from other vaccine trials or epidemiological studies that provide information, for example, on how the risk of infection or probability of disease progression depend on covariates (such as age, ethnicity, sex, etc.). In the absence of prior knowledge (as in the analysis presented here), non-informative priors can be used, and the Bayesian posterior probabilities will be equivalent to the model likelihood as derived in maximum likelihood (frequentist) approaches. Finally, Rstan provides an open-source, computationally efficient platform in which code is simple and straightforward to edit, allowing users to extend models of infection and disease to their specific scenario of interest.

In conclusion, JAMOVET is a powerful tool for evaluating vaccine trials against infections with multiple dependent outcomes and identifying the stages in the disease process at which the vaccine acts. It can be used to correct for known biases and identify predictors of key outcomes, enabling more targeted vaccine development and distribution.

## Data Availability

Anonymised participant data is available upon requests directed to the Oxford Vaccine Group*. Proposals will be reviewed and approved by the sponsor, investigator, and collaborators on the basis of scientific merit. After approval of a
proposal, data can be shared through a secure online platform after signing a data access agreement. All data will be made available for a
minimum of 5 years from the end of the trial.
*Prof Andrew J Pollard, Oxford Vaccine Group, Department of Paediatrics, University of Oxford,
Oxford OX3 7LE, UK andrew.pollard@paediatrics.
ox.ac.uk.

## References

1. Halloran ME, Longini IM, Jr., Struchiner CJ. Estimability and interpretation of vaccine efficacy using frailty mixing models. Am J Epidemiol. Jul 1 1996;144(1):83–97. doi:10.1093/oxfordjournals.aje.a008858

2. Kahn R, Hitchings M, Bellan S, Lipsitch M. Impact of stochastically generated heterogeneity in hazard rates on individually randomized vaccine efficacy trials. Clin Trials. Apr 2018;15(2):207–211. doi:10.1177/1740774517752671

3. Gomes MGM, Gordon SB, Lalloo DG. Clinical trials: The mathematics of falling vaccine efficacy with rising disease incidence. Vaccine. Jun 8 2016;34(27):3007–3009. doi:10.1016/j.vaccine.2016.04.065

4. Kahn R, Schrag SJ, Verani JR, Lipsitch M. Identifying and Alleviating Bias Due to Differential Depletion of Susceptible People in Postmarketing Evaluations of COVID-19 Vaccines. Am J Epidemiol. Mar 24 2022;191(5):800–811. doi:10.1093/aje/kwac015

5. Halloran ME, Auranen K, Baird S, et al. Simulations for designing and interpreting intervention trials in infectious diseases. BMC Med. Dec 29 2017;15(1):223. doi:10.1186/s12916-017-0985-3

6. Dean NE, Longini IM. The ring vaccination trial design for the estimation of vaccine efficacy and effectiveness during infectious disease outbreaks. Clin Trials. Aug 2022;19(4):402–406. doi:10.1177/17407745211073594

7. Jonesteller CL, Burnett E, Yen C, Tate JE, Parashar UD. Effectiveness of Rotavirus Vaccination: A Systematic Review of the First Decade of Global Postlicensure Data, 2006-2016. Clin Infect Dis. Sep 1 2017;65(5):840–850. doi:10.1093/cid/cix369

8. What defines an efficacious COVID-19 vaccine? A review of the challenges assessing the clinical efficacy of vaccines against SARS-CoV-2, 21 e26–e35 (2021).

9. Overview of Vaccine Effects and Study Designs, 19–45 (Springer-Verlag New York 2010).

10. Williams LR, Ferguson NM, Donnelly CA, Grassly NC. Measuring vaccine efficacy against infection and disease in clinical trials: sources and magnitude of bias in COVID-19 vaccine efficacy estimates. Clin Infect Dis. Oct 26 2021;doi:10.1093/cid/ciab914

11. Lachenbruch PA. Sensitivity, specificity, and vaccine efficacy. Controlled Clinical Trials 1998. p. 569–574.

12. Jackson ML, Rothman KJ. Effects of imperfect test sensitivity and specificity on observational studies of influenza vaccine effectiveness. Vaccine. Mar 10 2015;33(11):1313–6. doi:10.1016/j.vaccine.2015.01.069

13. Aalen OO, Johansen S. An Empirical Transition Matrix for Non-Homogeneous Markov Chains Based on Censored Observations. Scandinavian Journal of Statistics. 1978;5:141–150.

14. Le-Rademacher JG, Therneau TM, Ou FS. The Utility of Multistate Models: A Flexible Framework for Time-to-Event Data. Curr Epidemiol Rep. 2022;9(3):183–189. doi:10.1007/s40471-022-00291-y

15. Hougaard P. Multi-state models: a review. Lifetime Data Anal. Sep 1999;5(3):239–64. doi:10.1023/a:1009672031531

16. Kang M, Lagakos SW. Evaluating the role of human papillomavirus vaccine in cervical cancer prevention. Stat Methods Med Res. Apr 2004;13(2):139–55. doi:10.1191/0962280204sm358ra

17. O’Connor AM. Interpretation of odds and risk ratios. J Vet Intern Med. May-Jun 2013;27(3):600–3. doi:10.1111/jvim.12057

18. Williams LR, Ferguson NM, Donnelly CA, Grassly NC. Measuring Vaccine Efficacy Against Infection and Disease in Clinical Trials: Sources and Magnitude of Bias in Coronavirus Disease 2019 (COVID-19) Vaccine Efficacy Estimates. Clin Infect Dis. Aug 24 2022;75(1):e764–e773. doi:10.1093/cid/ciab914

19. 1. Barrett JR, Belij-Rammerstorfer S, Dold C, et al. Phase 1/2 trial of SARS-CoV-2 vaccine ChAdOx1 nCoV-19 with a booster dose induces multifunctional antibody responses. Nat Med. Feb 2021;27(2):279–288. doi:10.1038/s41591-020-01179-4

20. Folegatti PM, Ewer KJ, Aley PK, et al. Safety and immunogenicity of the ChAdOx1 nCoV-19 vaccine against SARS-CoV-2: a preliminary report of a phase 1/2, single-blind, randomised controlled trial. Lancet. Aug 15 2020;396(10249):467–478. doi:10.1016/S0140-6736(20)31604-4

21. Ramasamy MN, Minassian AM, Ewer KJ, et al. Safety and immunogenicity of ChAdOx1 nCoV-19 vaccine administered in a prime-boost regimen in young and old adults (COV002): a single-blind, randomised, controlled, phase 2/3 trial. Lancet. Dec 19 2021;396(10267):1979–1993. doi:10.1016/S0140-6736(20)32466-1

22. Voysey M, Clemens SAC, Madhi SA, et al. Safety and efficacy of the ChAdOx1 nCoV-19 vaccine (AZD1222) against SARS-CoV-2: an interim analysis of four randomised controlled trials in Brazil, South Africa, and the UK. Lancet. Jan 9 2021;397(10269):99–111. doi:10.1016/S0140-6736(20)32661-1

23. Skittrall JP, Wilson M, Smielewska AA, et al. Specificity and positive predictive value of SARS-CoV-2 nucleic acid amplification testing in a low-prevalence setting. Clin Microbiol Infect. Mar 2021;27(3):469 e9-469 e15. doi:10.1016/j.cmi.2020.10.003

24. Hellewell J, Russell TW, Investigators S, et al. Estimating the effectiveness of routine asymptomatic PCR testing at different frequencies for the detection of SARS-CoV-2 infections. BMC Med. Apr 27 2021;19(1):106. doi:10.1186/s12916-021-01982-x

25. Halloran ME, Longini IM, Struchiner CJ. Design and analysis of vaccine studies. Overview of vaccine effects and study designs. Springer-Verlag; 2010:19–45.

26. loo: Efficient leave-one-out cross-validation and WAIC for Bayesian models. Version 2.5.1. 2022. https://mc-stan.org/loo/

27. RStan: the R interface to Stan. 2020.

28. R Core Team. R: A Language and Environment for Statistical Computing. https://www.R-project.org/

29. Gelman A, Jakulin A, Pittau MG, Su Y-S. A weakly informative default prior distribution for logistic and other regression models. The Annals of Applied Statistics. 2008;2(4):1360–1383, 24.

30. Emary KRW, Golubchik T, Aley PK, et al. Efficacy of ChAdOx1 nCoV-19 (AZD1222) vaccine against SARS-CoV-2 variant of concern 202012/01 (B.1.1.7): an exploratory analysis of a randomised controlled trial. The Lancet 2021. p. 1351–1362.

31. Skittrall JP, Wilson M, Smielewska AA, et al. Specificity and positive predictive value of SARS-CoV-2 nucleic acid amplification testing in a low-prevalence setting. Clinical Microbiology and Infection: Elsevier B.V.; 2021. p. 469.e9-469.e15.

32. Braunstein GD, Schwartz L, Hymel P, Fielding J. False Positive Results With SARS-CoV-2 RT-PCR Tests and How to Evaluate a RT-PCR-Positive Test for the Possibility of a False Positive Result. J Occup Environ Med. Mar 1 2021;63(3):e159–e162. doi:10.1097/JOM.0000000000002138

33. Menni C, Klaser K, May A, et al. Vaccine side-effects and SARS-CoV-2 infection after vaccination in users of the COVID Symptom Study app in the UK: a prospective observational study. Lancet Infect Dis. Jul 2021;21(7):939–949. doi:10.1016/S1473-3099(21)00224-3

34. Antonelli M, Penfold RS, Merino J, et al. Risk factors and disease profile of post-vaccination SARS-CoV-2 infection in UK users of the COVID Symptom Study app: a prospective, community-based, nested, case-control study. Lancet Infect Dis. Jan 2022;22(1):43–55. doi:10.1016/S1473-3099(21)00460-6

35. Fulop T, Larbi A, Dupuis G, et al. Immunosenescence and Inflamm-Aging As Two Sides of the Same Coin: Friends or Foes? Front Immunol. 2017;8:1960. doi:10.3389/fimmu.2017.01960

36. Pereira B, Xu XN, Akbar AN. Targeting Inflammation and Immunosenescence to Improve Vaccine Responses in the Elderly. Front Immunol. 2020;11:583019. doi:10.3389/fimmu.2020.583019

37. Nguyen LH, Drew DA, Graham MS, et al. Risk of COVID-19 among front-line health-care workers and the general community: a prospective cohort study. Lancet Public Health. Sep 2020;5(9):e475–e483. doi:10.1016/S2468-2667(20)30164-X

38. Ward H, Atchison C, Whitaker M, et al. SARS-CoV-2 antibody prevalence in England following the first peak of the pandemic. Nat Commun. Feb 10 2021;12(1):905. doi:10.1038/s41467-021-21237-w

39. Kompaniyets L, Goodman AB, Belay B, et al. Body Mass Index and Risk for COVID-19-Related Hospitalization, Intensive Care Unit Admission, Invasive Mechanical Ventilation, and Death - United States, March-December 2020. MMWR Morb Mortal Wkly Rep. Mar 12 2021;70(10):355–361. doi:10.15585/mmwr.mm7010e4

40. Mathur R, Rentsch CT, Morton CE, et al. Ethnic differences in SARS-CoV-2 infection and COVID-19-related hospitalisation, intensive care unit admission, and death in 17 million adults in England: an observational cohort study using the OpenSAFELY platform. Lancet. May 8 2021;397(10286):1711–1724. doi:10.1016/S0140-6736(21)00634-6

41. Damme WV, Dahake R, van de Pas R, Vanham G, Assefa Y. COVID-19: Does the infectious inoculum dose-response relationship contribute to understanding heterogeneity in disease severity and transmission dynamics? Medical Hypotheses. 2021;146

42. Schrock JM, Ryan DT, Saber R, et al. Cohabitation With a Known Coronavirus Disease 2019 Case Is Associated With Greater Antibody Concentration and Symptom Severity in a Community-Based Sample of Seropositive Adults. Open Forum Infect Dis. Jul 2021;8(7):ofab244. doi:10.1093/ofid/ofab244

