## Supplementary materials for "A novel approach for estimating vaccine efficacy for infections with multiple outcomes: application to a COVID-19 vaccine trial"

#### Supplementary Methods

**Model extension: Additional stages of severity**

The model can be extended to account for additional stages of severity, such as severe symptoms, hospitalisation and death. Consider a disease with outcomes classified into three categories, C1, C2 and C3, representing, for example, asymptomatic infection, mild symptomatic infection and severe symptomatic infection, respectively. The risk of infection is given by $\Lambda$ , the probability of developing symptoms, given infection, $\theta_{C1\rightarrow C2}$, and the probability of developing severe symptoms, given by $\theta_{C2\rightarrow C3}$. Each of these parameters can be written as a function of vaccination status, as with the simpler model with only two potential outcomes. A vaccine with protective effects against each of these stages: VE against infection ($VE_{in}$ ) > 0, VE against progression from C1 to C2 ($VE_{C1\rightarrow C2}$) > 0, and VE against progression from C2 to C3 ($VE_{C2\rightarrow C3}$) > 0. The cumulative number of C1 infections is given by

$$C1_i \sim \text{Poisson}\left(\Lambda_\text{i}\left(1-\theta_{C1\rightarrow C2,i}\right)\right)$$

where $\Lambda_\text{i}$ represents the cumulative risk of infection and $\theta_{C1\rightarrow C2}$ represents the probability of progressing to symptoms, given infection, for an individual of type i. The cumulative number of C2 infections is given by

$$C2_i \sim \text{Poisson}\left(\Lambda_\text{i}\theta_{C1\rightarrow C2,i}\left(1-\theta_{C2\rightarrow C3,i}\right)\right)$$

and C3 can be described by

$$C3_i \sim \text{Poisson}\left(\Lambda_\text{i}\theta_{C1\rightarrow C2,i}\theta_{C2\rightarrow C3,i}\right)$$

Like with the simple model in which infections are classified by a binary symptom status, the probability of progressing from one stage of infection to another can be estimated using logistic regression. The model can be extended to any number of additional stages or alternative disease pathways as for standard multistate models.

**Model extension: Additional covariates**

The model can be extended to model covariates on the force of infection, probability of symptoms, $VE_{in}$ and $VE_{pr}$.

$$\log\left(\Lambda_i\right) = \alpha + \beta \times vaccinated_i + \boldsymbol{\theta} \times \boldsymbol{X_i} + \boldsymbol{\vartheta} \times vaccinated_i \times \boldsymbol{X_i} + \log(t_i)$$

$$\text{logit}(\text{s}_i) = \gamma + \delta \times vaccinated_i + \boldsymbol{\varepsilon} \times \boldsymbol{X_i} + \boldsymbol{\pi} \times vaccinated_i \times \boldsymbol{X_i}$$

Where $\boldsymbol{\theta}$, $\boldsymbol{\varepsilon}$, $\boldsymbol{\vartheta}$ and $\boldsymbol{\pi}$ represent vectors of regression coefficients for the covariates $\boldsymbol{X}$.

**Estimating probability of asymptomatic infection detection in COV002**

We calculated individual-level probabilities of asymptomatic infection detection using COV002 swabbing data and data from Hellewell *et al.* (2021) [1] on PCR test sensitivity to asymptomatic

SARS-CoV-2 infections. We used the data on PCR sensitivity to asymptomatic infections over the course of infection (Supplementary Table 4) to estimate the probability of asymptomatic infection detection for different frequencies of testing.

For example, for weekly testing, accounting for the possibility that a weekly test could be taken on any day in the first week after infection, the probability of asymptomatic infection detection was 83.3% (Supplementary Table 5). This was the maximum sensitivity value, used for participants who retuned all expected weekly tests during their follow up.

To estimate the sensitivity of asymptomatic infection detection in each individual, we estimated the proportion of expected weekly asymptomatic tests they returned. We made a simplifying assumption that all tests were taken at a regular frequency to convert this to an average frequency of testing. We then converted this to the probability of asymptomatic infection detection using the same process as for participants who tested weekly.

### Supplementary Tables

Supplementary Table 1. Number of true and false positives for asymptomatic infection in each simulated dataset scenario

| | Scenario 1<br>Specificity and asymptomatic sensitivity = 100% | Scenario 2<br>Specificity = 99.9% | Scenario 3<br>Asymptomatic sensitivity = 50% | Scenario 4<br>Asymptomatic adherence = 50% and specificity = 99.9% |
|---|---|---|---|---|
| N false positives | 0 (0-0) | 488 (444-529) | 0 (0-0) | 249 (220-279) |
| Vaccine | 0 (0-0) | 247 (216-277) | 0 (0-0) | 126 (105-147) |
| Placebo | 0 (0-0) | 241 (213-272) | 0 (0-0) | 123 (103-144) |
| N true positives | 719 (669-770) | 700 (652-753) | 545 (499-586) | 538 (494-583) |
| Vaccine | 244 (214-274) | 238 (208-268) | 160 (138-184) | 157 (133-181) |
| Placebo | 475 (436-516) | 464 (422-505) | 384 (345-422) | 380 (345-417) |
| PPV | 100 (100-100) | 58.9 (56.3-61.7) | 100 (100-100) | 68.4 (65.1-71.7) |
| Vaccine | 100 (100-100) | 49.0 (44.5-53.4) | 100 (100-100) | 55.6 (50.0-61.3) |
| Placebo | 100 (100-100) | 65.9 (62.2-69.3) | 100 (100-100) | 75.6 (71.7-79.2) |

PPV = Positive predictive value

All values are the mean and 95$^{th}$ percentiles of 1000 simulations.

Supplementary Table 2. Classification of candidate variables

| Candidate variable | Variable type(s) |
|---|---|
| **Age** | • Continuous<br>• Categorical (10 yr cats)<br>• Binary (+/-55 yrs and +/-65 yrs) |
| **Sex** | • Binary |
| **BMI*** | • Continuous<br>• Categorical (underweight, healthy weight, overweight, obese)<br>• Binary (obese y/n and overweight or obese y/n) |
| **Ethnicity** | • Categorical (white, black, Asian, prefer not to say)<br>• Binary (white y/n) |
| **HCW status** | • Categorical (HCW no COVID patients, HCW 1+ COVID patients, non HCW)<br>• Binary (HCW y/n) |
| **Comorbidities** | • Binary (comorbidity y/n)<br>• Binary (diabetes y/n)<br>• Binary (respiratory disease y/n)<br>• Binary (CV disease y/n) |

*BMI: <18.5= underweight; ≥18.5 and <25 = healthy weight; ≥25 and <30 = overweight, ≥30 = obese.

Supplementary Table 3. SARS-CoV-2 infections by severity in COV002.

| Trial Arm | Asymptomatic[1] | Mild-Moderate symptoms | Severe Symptoms[2] | Total |
|---|---|---|---|---|
| **Vaccine** | 129 | 67 | 0 | 196 |
| **Placebo** | 162 | 229 | 2 | 393 |

[1] Primary case definition

[2] Defined as a WHO clinical progression score ≥ 6

Supplementary Table 4. PCR test sensitivity to asymptomatic infection detection, estimated in Hellewell et al [1].

| Days since infection | Median | Lower | Upper |
|---|---|---|---|
| 0 | 0.005 | 0.000 | 0.020 |
| 1 | 0.039 | 0.005 | 0.161 |
| 2 | 0.246 | 0.048 | 0.766 |
| 3 | 0.710 | 0.226 | 0.881 |
| 4 | 0.785 | 0.559 | 0.892 |
| 5 | 0.767 | 0.613 | 0.885 |
| 6 | 0.725 | 0.574 | 0.857 |
| 7 | 0.677 | 0.529 | 0.818 |
| 8 | 0.626 | 0.484 | 0.771 |
| 9 | 0.571 | 0.437 | 0.720 |
| 10 | 0.515 | 0.390 | 0.660 |
| 11 | 0.459 | 0.344 | 0.597 |
| 12 | 0.404 | 0.296 | 0.534 |
| 13 | 0.351 | 0.252 | 0.469 |
| 14 | 0.302 | 0.211 | 0.411 |
| 15 | 0.257 | 0.170 | 0.357 |
| 16 | 0.217 | 0.137 | 0.311 |
| 17 | 0.180 | 0.108 | 0.268 |
| 18 | 0.149 | 0.083 | 0.229 |
| 19 | 0.122 | 0.065 | 0.198 |
| 20 | 0.100 | 0.050 | 0.168 |
| 21 | 0.082 | 0.038 | 0.144 |
| 22 | 0.066 | 0.028 | 0.124 |
| 23 | 0.054 | 0.021 | 0.106 |
| 24 | 0.044 | 0.016 | 0.091 |
| 25 | 0.035 | 0.012 | 0.078 |
| 26 | 0.028 | 0.009 | 0.066 |
| 27 | 0.023 | 0.006 | 0.056 |
| 28 | 0.018 | 0.005 | 0.048 |
| 29 | 0.014 | 0.004 | 0.041 |
| 30 | 0.012 | 0.003 | 0.035 |

Supplementary Table 5. Calculation of the probability of asymptomatic infection detection for weekly PCR testing.

| First test (days since infection) | Equation | Median | Lower CI | Upper CI |
|---|---|---|---|---|
| Day 0 | =1-(1-0.005)*(1-0.677)*(1-0.302)*(1-0.082)*(1-0.018) | 0.798 | 0.645 | 0.915 |
| Day 1 | =1-(1-0.039)*(1-0.626)*(1-0.257)*(1-0.066)*(1-0.014) | 0.754 | 0.587 | 0.896 |
| Day 2 | =1-(1-0.246)*(1-0.571)*(1-0.217)*(1-0.054)*(1-0.012) | 0.763 | 0.549 | 0.961 |
| Day 3 | =1-(1-0.710)*(1-0.515)*(1-0.180)*(1-0.044) | 0.890 | 0.585 | 0.973 |
| Day 4 | =1-(1-0.785)*(1-0.459)*(1-0.149)*(1-0.035) | 0.905 | 0.738 | 0.969 |
| Day 5 | =1-(1-0.767)*(1-0.404)*(1-0.122)*(1-0.028) | 0.881 | 0.748 | 0.960 |
| Day 6 | =1-(1-0.725)*(1-0.351)*(1-0.100)*(1-0.023) | 0.843 | 0.699 | 0.940 |
| **Average** | | **0.833** | **0.650** | **0.945** |

#### References


1. Hellewell J, Russell TW, Investigators S, et al. Estimating the effectiveness of routine asymptomatic PCR testing at different frequencies for the detection of SARS-CoV-2 infections. BMC Med **2021**; 19(1): 106.
